# Social inequalities on Cancer Incidence and Mortality in a Brazilian City

**DOI:** 10.1101/2025.10.16.25338205

**Authors:** Andrea Paula Bruno von Zuben, Maria do Carmo Ferreira, Marilisa Berti de Azevedo Barros, Juliana Nativo, M. Elvira P. Correa, Carmino Antônio de Souza

## Abstract

**Background:** Cancer represents one of the leading global public health challenges, with its burden shaped not only by biological factors but also by social and economic inequalities. In Brazil, even municipalities with very high Human Development Index (HDI) show persistent disparities. This study assessed temporal changes in cancer incidence, mortality, and social inequalities in Campinas, São Paulo State, Brazil.

**Methods:** We conducted a cross-sectional study using data from the Population-Based Cancer Registry and the Mortality Information System for 2010–2014 and 2015–2019. Age-standardized incidence and mortality rates were estimated for the most common cancers in men and women, stratified by levels of social vulnerability based on the São Paulo Social Vulnerability Index. Inequalities were analyzed using rate ratios and the Relative Index of Inequality (RII).

**Results:** Among men, prostate and stomach cancer incidence declined, while mortality remained stable except for reductions in stomach cancer. Socially vulnerable men showed persistently higher mortality from prostate, stomach, and oral cavity cancers, with disparities widening over time. Among women, incidence increased for breast and lung cancers, and overall mortality rose, particularly from lung cancer. Vulnerable women exhibited consistently higher cervical cancer incidence and mortality, while breast cancer incidence remained higher in less vulnerable groups. Inequalities in colorectal cancer incidence and mortality narrowed over time for both sexes.

**Conclusion:** Despite Campinas’ high HDI and urbanization, significant social inequalities in cancer incidence and mortality persist, with worsening disparities for some cancer types. These findings highlight the need for equity-focused health policies to ensure timely diagnosis, treatment, and palliative care for socially vulnerable populations.

## Introduction

Neoplasms currently rank as the second leading cause of mortality in most countries, surpassed only by cardiovascular diseases. As deaths from acute myocardial infarction and stroke decline in some nations, cancer-related deaths have already overtaken cardiovascular mortality to become the primary cause of death. For premature deaths between ages 30 and 69, neoplasms constitute the leading cause in 57 countries^1^.

Cancer represents one of the greatest challenges to global public health. Its burden stems not only from biological factors but also from social and economic determinants. Cancer inequalities vary depending on the indicator analyzed—incidence or mortality— and evolve over time^2,3,4^.

In countries like Brazil, socioeconomic inequalities play a crucial role in cancer incidence, mortality, and access to oncological care, generating persistent disparities even in regions with high Human Development Index (HDI) scores. Campinas, a municipality in São Paulo State, Brazil, with a very high HDI (0.805) and extensive urbanization (98.28%), exhibits marked social disparities in health indicators, making it an important setting for investigating how social inequities manifest in cancer patterns^5,6^. International monitoring of cancer inequalities has shown that disparities remain unchanged or have even widened in some cases^4,7,8^.

The municipality has established a well-structured Population-Based Cancer Registry (PBCR) with high-quality information spanning 10 consecutive years, enabling analysis of changes in incidence and mortality across intra-urban social strata. Population-based registry studies are essential for supporting public policies and evaluating health intervention impacts. In Campinas, data availability from the PBCR and the Mortality Information System (MIS), combined with indicators such as the São Paulo Social Vulnerability Index (IPVS)^9^, enables stratified and georeferenced analyses that provide evidence on how social determinants influence cancer incidence and mortality.

This study aimed to assess changes in incidence, mortality, and social inequalities for the most common cancers in Campinas between 2010–2014 and 2015–2019, considering levels of social vulnerability.

By examining these patterns in a developed municipality still marked by internal disparities, this work contributes to the debate on the need for public policies targeting the most vulnerable populations to reduce inequities in access to early diagnosis, timely treatment, and quality palliative care.

## Material and Methods

### Study Design and Setting

This cross-sectional study evaluated incidence and mortality rates of the most frequent neoplasms among men and women residing in Campinas during 2010–2014 and 2015– 2019, before pandemic period. We stratified analyses by area of residence according to social vulnerability level.

Campinas is a large municipality located approximately 100 km from São Paulo, the state capital. On July 1, 2024, its estimated population reached 1,185,977 inhabitants according to the Brazilian Institute of Geography and Statistics (IBGE), placing Campinas as the second most populous municipality among non-state capitals. The 2022 Demographic Census recorded an urbanization rate of 98.28%. In 2010, Campinas achieved a Municipal Human Development Index (MHDI) of 0.805, classified as very high^10^.

## Data Sources and Cancer Types

We obtained new cancer cases from the Population-Based Cancer Registry and retrieved deaths from the Mortality Information System. Both systems are managed by the Campinas Municipal Health Department. The following neoplasm types were included with their corresponding codes from the International Statistical Classification of Disease and Related Health Problems (ICD-10):

**Male:** prostate (C61), colorectal (C18–C20), lung (C33–C34), stomach (C16), oral cavity and oropharynx (C00–C14), and total neoplasms excluding non-melanoma skin cancer (C00–C97, except C44). **Female**: breast (C50), cervix (C53), colorectal (C18–C20), lung (C33–C34), stomach (C16), and total neoplasms excluding non-melanoma skin cancer.

## Social Vulnerability Classification

We obtained population estimates by coverage area of Primary Health Units from IBGE data and the Campinas Municipal Health Department^11^. Social vulnerability strata (EVS) were defined based on the São Paulo Social Vulnerability Index (IPVS-2010), developed by the Fundação Sistema Estadual de Análise de Dados (Seade Foundation), a public institution linked to the Government of São Paulo State specializing in statistical and sociodemographic data production and analysis^9^.

Primary Health Units were classified according to the proportion of census tracts in each IPVS category within their coverage area, then grouped into three levels: EVS 1 (lowest vulnerability), EVS 2 (intermediate vulnerability), and EVS 3 (highest vulnerability) (Figure 1).

**Figure 1:**
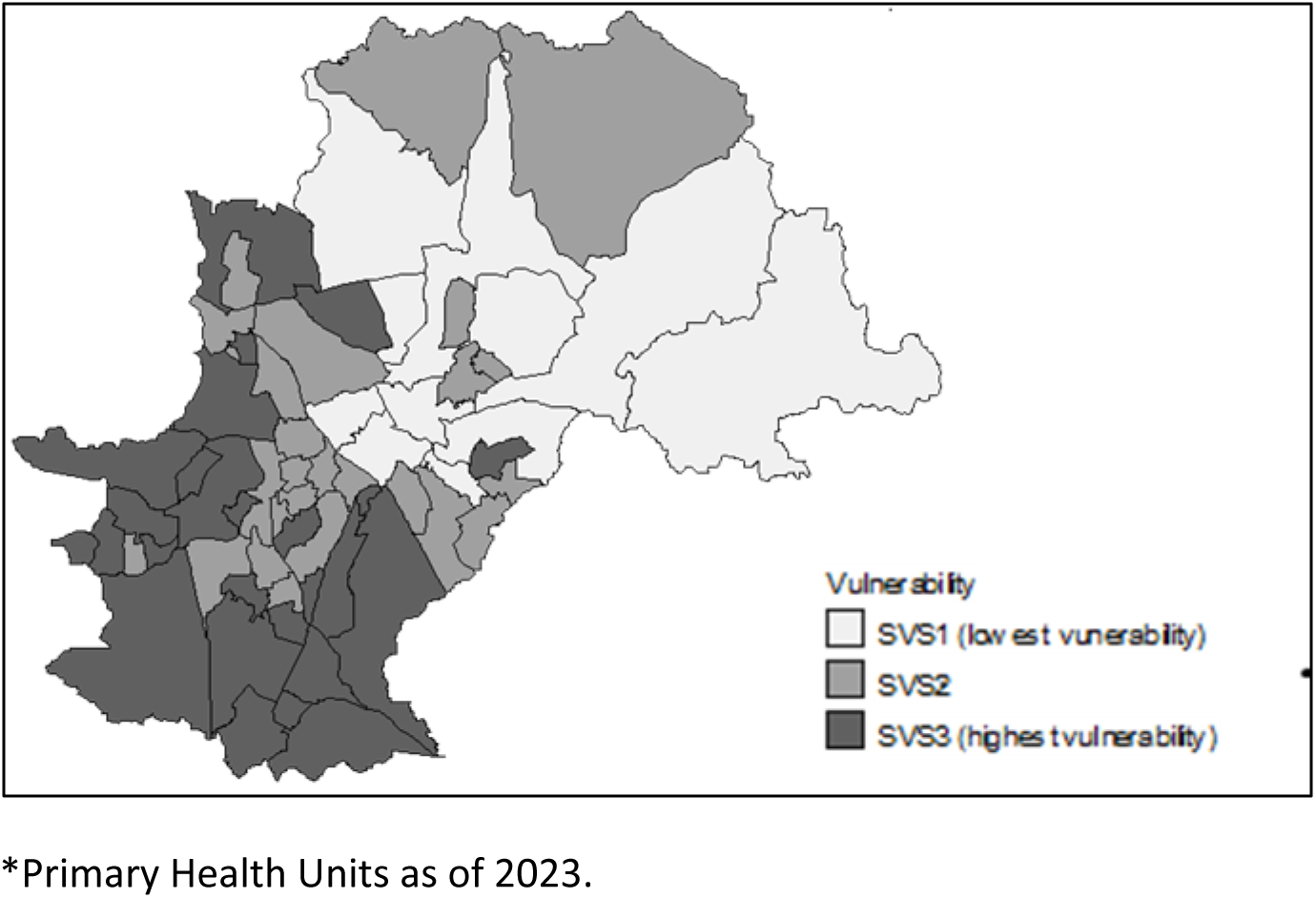
Coverage areas of Primary Health Units in the municipality of Campinas, according to social vulnerability strata. *\**.

## Statistical Analysis

We adjusted deaths from cervical cancer of unspecified portion (ICD-10: C55) and proportionally reallocated them between cervical cancer (C53) and uterine corpus cancer (C54) according to age group and vulnerability stratum for both study periods^12^. We age-standardized incidence and mortality rates using the direct method, with the 1960 world standard population as reference^13^.

Rate ratios (RRs) and their 95% confidence intervals (CIs) were estimated for incidence and mortality rates of each cancer type between 2010–2014 and 2015–2019. We also made comparisons between extreme EVS strata, contrasting the most vulnerable (EVS 3) with the least vulnerable (EVS 1). To measure inequalities in cancer incidence and mortality for total neoplasms and specific cancer types, we calculated the Relative Index of Inequality (RII)^14^.

For RII calculations, we used the RIIGEN module of Stata version 15.0 (StataCorp. LP) to generate a rank variable considering all social vulnerability strata. These were ordered from lowest to highest vulnerability and classified with a score from 0 to 1 based on cumulative relative population position. After analyzing linearity between health indicators of each stratum (dependent variable) and the rank (independent variable), we performed Poisson regression to estimate the RII value, which shows the relative difference between strata. Values greater than 1 indicate higher risk among groups with greater social vulnerability, while values less than 1 reveal higher risk among groups with lower vulnerability.

We performed statistical analyses using Microsoft Excel 2016® and Stata 15.0 (StataCorp, College Station, USA). The map of areas according to social vulnerability was created using TabWin Software, version 4.1.5. We adopted a significance level of 5% for all analyses.

## Results

### Overall Temporal Trends by Sex

Cancer incidence and mortality rates in Campinas across 2010–2014 and 2015–2019 revealed distinct patterns by sex. Among men, incidence rates fell for prostate cancer, RR = 0.94 (95% CI: 0.89–0.99), and stomach cancer, RR = 0.70 (95% CI: 0.72–0.93). Other neoplasms showed no statistically significant changes in incidence rates.

Regarding mortality during these same periods, rates declined only for stomach cancer, RR = 0.85 (95% CI: 0.72–0.99). Mortality rates for other cancer types remained stable.

Among women, incidence rates increased for breast cancer, RR = 1.14 (95% CI: 1.08– 1.20), and lung cancer, RR = 1.22 (95% CI: 1.07–1.40). Other female neoplasms showed no statistically significant changes in incidence rates. Concerning mortality, overall cancer mortality increased, RR = 1.06 (95% CI: 1.01–1.12), as did mortality from bronchus and lung cancer, RR = 1.25 (95% CI: 1.07–1.47). Other cancer types among women showed no statistically significant changes in mortality rates (Table 1).

**Table 1.** Age-standardized incidence and mortality rates of neoplasms, by sex and cancer type, in the periods 2010–2014 and 2015–2019. Campinas, Brazil.

| Primary site | Incidence rates per 100,000 population |  |  | Mortality rates per 100,000 population |  |  |
| --- | --- | --- | --- | --- | --- | --- |
|  | 2010–2014 | 2015–2019 | RR** | 2010–2014 | 2015–2019 | RR** |
| **Male** |  |  |  |  |  |  |
| All neoplasms* | 313.78<br>(307.19–<br>320.38) | 303.60 (297.27–<br>309.93) | 0.97 (0.94–<br>1.00) | 120.14 (116.10–<br>124.19) | 122.71 (118.74–<br>126.69) | 1.02 (0.87–<br>1.07) |
| Prostate | 97.41 (93.70–<br>101.12) | 91.53 (88.02–<br>95.04) | 0.94 (0.89–<br>0.99) | 11.55 (10.33–<br>12.77) | 12.18 (10.95–<br>13.40) | 1.05 (0.91–<br>1.22) |
| Colon and rectum | 34.39 (32.23–<br>36.56) | 36.30 (34.13–<br>38.47) | 1.06 (0.97–<br>1.15) | 12.41 (11.11–<br>13.70) | 14.27 (12.92–<br>15.63) | 1.15 (1.00–<br>1.32) |
| Bronchus and lung | 20.41 (18.74–<br>22.08) | 20.67 (19.01–<br>22.32) | 1.01 (0.90–<br>1.14) | 16.52 (15.02–<br>18.02) | 16.46 (14.99–<br>17.92) | 1.00 (0.88–<br>1.13) |
| Stomach | 18.23 (16.66–<br>19.80) | 14.92 (13.54–<br>16.30) | 0.70 (0.72–<br>0.93) | 11.53 (10.28–<br>12.77) | 9.77 (8.66–<br>10.89) | 0.85 (0.72–<br>0.99) |
| Oral cavity | 18.34 (16.76–<br>19.91) | 18.89 (17.32–<br>20.45) | 1.03 (0.91–<br>1.16) | 5.81 (4.92–6.69) | 6.04 (5.15–6.93) | 1.04 (0.84–<br>1.28) |
| **Female** |  |  |  |  |  |  |
| All neoplasms* | 242.97<br>(237.74–<br>248.20) | 242.73 (237.67–<br>247.79) | 1.00 (0.97–<br>1.03) | 76.74 (73.86–<br>79.61) | 81.52 (78.65–<br>84.38) | 1.06 (1.01–<br>1.12) |
| Breast | 72.45 (69.61–<br>75.28) | 82.39 (79.45–<br>85.33) | 1.14 (1.08–<br>1.20) | 13.52 (12.33–<br>14.72) | 14.33 (13.14–<br>15.53) | 1.06 (0.94–<br>1.20) |
| Cervix uteri | 7.08 (6.20–<br>7.96) | 7.37 (6.50–8.24) | 1.04 (0.88–<br>1.24) | 2.88 (2.32–3.44) | 2.96 (2.40–3.52) | 1.03 (0.78–<br>1.35) |
| Colon and rectum | 27.67 (25.94–<br>29.40) | 28.31 (26.61–<br>30.02) | 1.02 (0.94–<br>1.12) | 8.78 (7.83–9.74) | 9.62 (8.64–<br>10.59) | 1.10 (0.94–<br>1.27) |
| Bronchus and lung | 10.58 (9.50–<br>11.65) | 12.90 (11.74–<br>14.06) | 1.22 (1.07–<br>1.40) | 7.71 (6.80–8.62) | 9.67 (8.68–<br>10.67) | 1.25 (1.07–<br>1.47) |
| Stomach | 7.76 (6.86–<br>8.66) | 7.66 (6.79–8.54) | 0.99 (0.84–<br>1.14) | 3.94 (3.32–4.56) | 3.67 (3.08–4.26) | 0.93 (0.74–<br>1.17) |

Standardized rates based on the 1960 world standard population; \*Except C44 (non-melanoma skin cancer), \*\*p<0.05, \*p<0.05; \*\*Except C44 (non-melanoma skin cancer), \*p<0.05; \*Rates per 100,000, age-adjusted to the 1960 world standard population; \*\*Except C44 (non- melanoma skin cancer), \*p<0.05; \*Rates per 100,000, age-adjusted to the 1960 world standard population

## Inequality Assessment Across Social Strata

The evaluation of inequality magnitude in incidence and mortality used both rate ratios between extreme strata (EVS 3/EVS 1) and the Relative Index of Inequality. These approaches yielded similar results, with some differences in statistical significance.

## Men’s Cancer Patterns

Among men, incidence of total neoplasms and colorectal cancer was significantly lower among the most vulnerable groups according to both measures in both periods analyzed. We detected no inequality in prostate cancer incidence during either period with both indicators. Bronchus and lung cancer incidence, which was lower in the most socially vulnerable group in 2010–2014, showed no differences in 2015–2019 according to either indicator. Conversely, for stomach and oral cavity cancers, incidence rates were elevated among men in the most vulnerable stratum during both study periods.

Concerning mortality patterns, prostate and oral cavity cancer rates were higher among men in the most vulnerable group in 2010–2014. In the subsequent period, inequality widened dramatically, reaching three times the rate recorded among the least vulnerable. For total neoplasms and bronchus and lung cancer, we detected no mortality inequalities in 2010–2014. Yet in 2015–2019, mortality was greater among men in the most vulnerable stratum according to both indicators for all neoplasms, and according to RII for bronchus and lung cancer.

For colorectal cancer, mortality was lower among men in the most vulnerable group in 2010–2014, with no inequality detected in 2015–2019. Among socially vulnerable men, stomach cancer mortality in both periods, as measured by the RII, exceeded that in the least vulnerable group (Table 2).

**Table 2.** Social inequalities in cancer incidence and mortality by sex, in the periods 2010– 2014 and 2015–2019. Campinas/SP.

| Primary site | Incidence RR (strata 3 vs. 1 of Social Vulnerability) |  |  |  | Mortality RR (strata 3 vs. 1 of Social Vulnerability) |  |  |  |
| --- | --- | --- | --- | --- | --- | --- | --- | --- |
|  | 2010-2014 |  | 2015-2019 |  | 2010-2014 |  | 2015-2019 |  |
| **Male** | RR 3/1** | IRD* | RR 3/1** | IRD* | RR3/1** | IRD* | RR 3/1** | IRD* |
| All neoplasms* | 0.74<br>0.70-0.78 | 0.64*<br>0.63-0.65 | 0.87<br>0.82-0.92 | 0.81*<br>0.72-0.93 | 1.07<br>0.98-1.18 | 1.11<br>0.96-1.28 | 1.28<br>1.17-1.40 | 1.40*<br>1.13-1.72 |
| Prostate | 0.96<br>0.87;1.07 | 0.94<br>0.89-1.00 | 1.01<br>0.91;1.12 | 1.01<br>0.88-1.15 | 1.50<br>1.10-2.04 | 2.20*<br>1.72-2.82 | 2.33<br>1.72-3.15 | 3.16*<br>2.37-4.20 |
| Colon and rectum | 0.44<br>0.37;0.53 | 0.31*<br>0.22-0.44 | 0.67<br>0.57-0.78 | 0.57*<br>0.46-0.69 | 0.61<br>0.45-0.82 | 0.53*<br>0.30-0.94 | 0.89<br>0.69-1.15 | 0.85<br>0.65-1.11 |
| Bronchus and lung | 0.79<br>0.63;0.99 | 0.71*<br>0.69-0.74 | 1.00<br>0.80;1.24 | 0.96<br>0.90-1.04 | 1.00<br>0.77-1.28 | 1.00<br>0.89-1.12 | 1.16<br>0.91-1.48 | 1.24*<br>1.17-1.31 |
| Stomach | 1.26<br>1.00;1.60 | 1.41*<br>1.35-1.47 | 1.46<br>1.13;1.88 | 1.70*<br>1.50-1.92 | 1.43<br>0.92-2.22 | 2.38*<br>2.01-2.80 | 1.62<br>0.99-2.67 | 2.08*<br>1.70-2.54 |
| Oral cavity | 1.42<br>1.13-1.78 | 1.41*<br>1.35-1.47 | 1.35<br>1.09-1.68 | 1.70*<br>1.51-1.92 | 1.86<br>1.19-2.89 | 2.32*<br>1.61-3.34 | 2.44<br>1.61-3.72 | 3.50*<br>3.02-4.05 |
| **Female** | 2010-2014 |  | 2015-2019 |  | 2010-2014 |  | 2015-2019 |  |
| All neoplasms* | 0.64<br>0.60-0.68 | 0.52*<br>0.49-0.54 | 0.78<br>0.74-0.82 | 0.69*<br>0.65-0.74 | 0.94<br>0.85-1.04 | 1.09<br>0.87-1.34 | 1.17<br>1.06-1.29 | 1.24*<br>1.04-1.49 |
| Breast | 0.52<br>0.47-0.58 | 0.39*<br>0.36-0.41 | 0.58<br>0.53-0.64 | 0.45*<br>0.39-0.53 | 0.68<br>0.53-0.88 | 0.69<br>0.46;1.05 | 0.75<br>0.59-0.94 | 0.66*<br>0.52-0.84 |
| Cervix uteri | 2.30<br>1.62-3.23 | 3.52*<br>3.12-3.97 | 2.05<br>1.49-2.82 | 2.99*<br>2.49-3.58 | 3.03<br>1.58-5.80 | 4.19*<br>3.96-4.42 | 2.48<br>1.42-4.34 | 4.01*<br>2.68-5.99 |
| Colon and rectum | 0.57<br>0.48-0.68 | 0.47*<br>0.32-0.68 | 0.98<br>0.83-1.16 | 0.97<br>0.84-1.12 | 0.69<br>0.59-0.81 | 0.72<br>0.51-1.01 | 1.18<br>0.89-1.57 | 1.27<br>0.99-1.63 |
| Bronchus and lung | 1.11<br>0.84-1.47 | 1.17<br>0.99-1.38 | 0.80<br>0.63-1.02 | 0.73*<br>0.70-0.76 | 1.16<br>0.85-1.60 | 1.45*<br>1.23-1.71 | 1.14<br>0.86-1.52 | 1.21*<br>1.02-1.43 |
| Stomach | 0.92<br>0.66-1.27 | 0.88*<br>0.87-0.89 | 1.42<br>1.09-1.86 | 1.69*<br>1.59-1.81 | 1.43<br>0.92-2.22 | 2.22*<br>2.21-2.24 | 1.62<br>0.99-2.67 | 1.83<br>0.85-3.96 |

\*RII - Relative Index of Inequality, $p < 0.05$ ; \*\*RR - Rate Ratio SES 3/1, 95% Confidence Interval

## Women’s Cancer Patterns

Among women, breast cancer and total neoplasm incidence was higher in the least vulnerable group during both periods. Still, the magnitude of this difference decreased over time, as indicated by both inequality measures. Cervical cancer incidence was consistently elevated among the most vulnerable women in both periods.

For colorectal cancer, incidence was higher in the least vulnerable women in 2010–2014, but this inequality disappeared in the subsequent period. Bronchus and lung cancer incidence showed no inequality in 2010–2014. Still, in 2015–2019, lower incidence was evident among women in the most vulnerable group according to the RII. For stomach cancer, incidence was lower among women in the most vulnerable group according to the RII in 2010–2014, with no inequality detected by the rate ratio. Yet in 2015–2019, incidence became higher in this same group, detected by both measures.

Regarding mortality, women in the most vulnerable group had higher cervical cancer mortality in both periods, as indicated by both measures. For total neoplasms, no mortality inequality was evident in 2010–2014. Yet in 2015–2019, mortality was greater among the most vulnerable women. For breast cancer, mortality remained higher among the least vulnerable women according to the rate ratio in both periods. On the other hand, the RII did not detect this inequality in 2010–2014 and showed lower mortality among the most vulnerable in 2015–2019.

For colorectal cancer, mortality was lower among women in the most vulnerable group by the rate ratio in 2010–2014, with no statistical significance by the RII. In 2015–2019, neither indicator detected statistically significant mortality differences. For bronchus and lung cancer, mortality was higher among women in the most vulnerable group in both periods according to the RII, but differences did not reach statistical significance by the rate ratio. For stomach cancer, higher mortality was evident among women in the most vulnerable group in 2010–2014 according to the RII, but in 2015–2019 the rate was not statistically significant.

## Discussion

### Context of Findings

The results from Campinas during 2010–2019 corroborate findings from national and international studies demonstrating social disparities in cancer incidence and mortality^7,16,17,18,19^. Despite its high Human Development Index and degree of urbanization, the municipality exhibits significant differences in indicators across population strata with varying levels of social vulnerability. This pattern underscores the persistence of socioeconomic inequalities in cancer over time, suggesting that economic growth alone is insufficient to guarantee equity in health outcomes^18^.

## Prostate Cancer: Declining Incidence, Widening Mortality Gaps

Prostate cancer demonstrates distinct trends across different world regions, influenced by factors such as screening, diagnostic access, and social inequalities. While Wang et al. (2022) documented increased incidence in 44 countries between 2015 and 2019^20^, data from Campinas indicate a decline during the same period, RR 0.94 (95% CI: 0.89– 0.99). This pattern aligns with national evidence indicating declining incidence in areas with greater socioeconomic development and in municipalities with high HDI in Mato Grosso^20^. The similarity with findings from England, where incidence also declined in wealthier regions, suggests that access to early diagnosis may strongly influence these trends^21^.

Although prostate cancer mortality remained stable in Campinas during the study period, social inequalities increased substantially. There was a threefold elevated risk of death among the most vulnerable men in 2015–2019 compared to the previous five-year period. This pattern reinforces findings by Costa et al.^22^ showing that improvements in social welfare reduce mortality risk, though this benefit is four times greater for white men compared to Black men.

Internationally, mortality reduction has been attributed to therapeutic advances in 32 countries^19^, although the magnitude of this decline has lessened in recent years. In contrast, South Korea has demonstrated that public policies aimed at health equity can reduce regional disparities in prostate cancer outcomes^23^. This highlights the absence of similar policies as a possible explanation for the worsening inequalities in Campinas.

## Stomach Cancer: Complex Gender-Specific Patterns

The reduction in stomach cancer incidence among men in this study aligns with the decline documented throughout Brazil. Azevedo e Silva et al.^24^, although focused on mortality, reinforces this trend, highlighting the sharp fall in mortality rates for stomach cancer across much of the country. This pattern suggests that socioeconomic disparities remain a key factor in the magnitude of decline in stomach cancer incidence in Brazil, attributable to differences in exposure to risk factors as well as access to early diagnosis.

Data from Campinas between 2010–2014 and 2015–2019 showed a reduction in both incidence (RR = 0.70; 95% CI: 0.72–0.93) and mortality (RR = 0.85; 95% CI: 0.72–0.99) for stomach cancer in men. Socially vulnerable men consistently had the highest incidence and mortality rates in both periods, indicating persistent inequities.

Among women, a reversal emerged: while incidence was lower in the most vulnerable group in 2010–2014, it became higher in 2015–2019. Increases were detected by both rate ratio and RII, suggesting worsening inequalities among women as well. These findings are consistent with national studies, such as that conducted in Greater Cuiabá^25^, which identified declining incidence of gastric cancer in men, particularly the elderly, and in adult and elderly women.

A population-based study conducted in South Korea between 1999 and 2018 showed a reduction in regional disparities in stomach cancer incidence in both men and women, despite overall high incidence^23^. The comparison with Campinas suggests that systematically implemented equity-oriented health policies may contribute to mitigating disparities in gastric cancer.

## Breast Cancer: The Epidemiological Transition

Data from Campinas reveal an increase in breast cancer incidence (RR = 1.14; 95% CI: 1.08–1.20) between 2010 and 2019. This pattern aligns with trends documented in developing countries such as China and South Korea, where incidence and mortality rates increased sharply between 2000 and 2015^26^. This scenario contrasts with reductions seen in developed nations such as the United States and the United Kingdom over the same period, illustrating the dynamic nature of the breast cancer epidemiological transition^27^.

Stratified analysis by social vulnerability in Campinas shows that incidence was consistently higher among women with lower vulnerability—a characteristic typical of countries with high HDI. This phenomenon suggests a “layered epidemiological transition,” in which different socioeconomic groups are at distinct stages of the breast cancer development curve. This pattern is similar to that documented in contexts such as India, where breast cancer has emerged as the leading female neoplasm^28^.

Regarding mortality, higher rates were evident among less vulnerable women according to rate ratio, but with a trend toward reduction among more vulnerable women when analyzed by RII in 2015–2019. This complexity emphasizes the urgency of differentiated policies, including implementation of organized screening programs, guaranteed equitable access to innovative treatments, and strengthening of information systems^29^.

## Lung Cancer: Shifting Gender Patterns

For bronchus and lung cancer, both incidence and mortality increased significantly in Campinas, with overall mortality rising (RR = 1.25; 95% CI: 1.07–1.47). This increase was particularly pronounced among women, the group in which incidence also grew substantially. Among men, incidence was lower in the most vulnerable group in 2010– 2014, but this difference disappeared in 2015–2019. Mortality, in turn, was elevated among men in the most vulnerable group in the latter period. Meanwhile, among women, inequalities persisted, with higher mortality in the most vulnerable group in both periods according to RII.

These findings align with national and international literature pointing to increasing burden among women and declining burden among men, primarily linked to smoking trends. International studies such as “Incidence and Mortality of Lung Cancer: Global Trends and Association with Socioeconomic Status”^30^ and “Distribution, Risk Factors, and Temporal Trends for Lung Cancer Incidence and Mortality: A Global Analysis”^31^ have identified consistent increases in incidence and mortality among women and declines among men, especially in high-income countries.

In Brazil, similar results were reported in temporal analyses of major cities. For instance, “Mortalidade por câncer nas capitais e no interior do Brasil: uma análise de quatro décadas”^24^ documented declining lung cancer mortality among men in the Southeast and South but increases among women. In Cuiabá, a local trends study suggested that social, economic, cultural, and biological factors influence these disparities, underscoring their complexity^32^.

Furthermore, Vaccarella et al. (2023) demonstrated that lung cancer is the main contributor to cancer mortality inequalities in Europe, accounting for 32% of deaths among men and 16% among women associated with educational inequalities^33^. In Brazil, regions with lower HDI presented greater increases or smaller reductions in mortality, as shown in “Inequalities in Lung Cancer Mortality Trends in Brazil, 2000–2015”^34^. A study on mortality in Campinas revealed that lung cancer primarily affects white men aged 70–79, with smoking as the main risk factor and lower educational attainment negatively influencing access to preventive measures^35^. Studies in the United States reinforce that local social vulnerability is a key predictor of cancer mortality^36^.

## Oral Cavity Cancer: Persistent and Widening Disparities

In both periods analyzed in Campinas, men in the most vulnerable stratum exhibited the highest incidence of oral cavity cancer, indicating a persistent inequality pattern in disease burden. According to the Global Oral Cancer Forum, incidence and mortality in Europe are strongly associated with smoking patterns, alcohol consumption, and unequal access to healthcare services^37^.

The intensification of inequalities in Campinas is particularly evident in oral cavity cancer mortality. The mortality ratio between the most and least vulnerable groups tripled from 2010–2014 to 2015–2019. Factors such as late diagnosis, restricted access to quality treatment, and limited access to information may explain part of this disparity. Schuurman et al. (2013) in the Netherlands showed that even in contexts of overall survival improvements, oral cavity cancer incidence and mortality remained high in more vulnerable subgroups, indicating that progress is not equally distributed^38^.

## Cervical Cancer: A Marker of Health Inequality

Consistently across both periods and indicators of incidence and mortality, women in the most vulnerable group in Campinas had the worst outcomes for cervical cancer. This highlights the close relationship between socioeconomic inequalities and women’s health. The study “Global Inequalities in Cervical Cancer Incidence and Mortality” (2018) showed that cervical cancer incidence and mortality rates are substantially higher in regions with low and medium HDI compared to those with high HDI^39^.

The global study reported a significant negative association between cervical cancer occurrence and all HDI components—including life expectancy, years of schooling, and gross national income. This underscores that cervical cancer is a major public health problem in lower-HDI countries. In Brazil, cervical cancer mortality has shown complex patterns. Although Cancer mortality in the Capitals and in the interior of Brazil: a four-decade analysis^24^ (1978–2017) indicated a general decline in cervical cancer mortality across most of the country, except in the interior of the North region, the study “Recent Changes in Cervical Cancer Mortality Trends in Southeastern Brazil” (1980–2020) raised specific concerns for São Paulo state^40^.

This study revealed a discrete but significant reversal in the declining trend of cervical cancer mortality in São Paulo beginning in 2014–2020, with a more pronounced increase among women aged 25–39. The authors highlighted that the consistent decline in Pap smear coverage within the Unified Health System since 2012 across all age groups may explain this trend, pointing to potential screening failures.

A previous study in Campinas identified a 57% decline in incidence and a 42% decline in cervical cancer mortality between 1991–1995 and 2010–2014, while incidence of in situ cervical carcinoma tripled in the same period^41^. These results suggest that early detection may have contributed to reduced incidence of invasive carcinoma and mortality. South Korean literature^42^ identified reductions in regional cervical cancer disparities among women, indicating that targeted interventions may mitigate these inequalities.

In Campinas, the persistence of cervical cancer disparities underscores the importance of strengthening organized screening programs that effectively reach the most vulnerable women and ensure proper follow-up of abnormal results, thereby reversing this trend and reducing the disease burden in this priority group.

## Colorectal Cancer: A Transitional Period

Colorectal cancer incidence and mortality rates for both sexes remained stable across the study periods. The highest incidence rates were recorded among men in the least vulnerable group in both periods, though at a smaller magnitude in 2015–2019 compared to 2010–2014. In 2010–2014, incidence among the least vulnerable men was 56% and 69% higher (rate ratio and RII, respectively) than in the most vulnerable. Yet in 2015–2019 this difference decreased to 39% (rate ratio) and 47% (RII).

Among women, incidence was also higher in the least vulnerable group in 2010–2014, but the difference diminished and even disappeared, according to RII, in 2015–2019. These results indicate a reduction in inequalities unfavorable to men and women in the most vulnerable stratum.

Regarding mortality, which in 2010–2014 was lower among men (by both indicators) and women (by rate ratio) in the most vulnerable group, no differences were detected in the most recent period. Even a reversal was evident among women, though not statistically significant, in 2015–2019. As with incidence, inequalities declined, reducing disadvantages for the most vulnerable.

These findings may indicate a transitional period, with incidence beginning to increase in the most vulnerable population. This situation has been documented in the United States and European countries in past decades^7,43^. In the U.S., between 1950 and 2013, colorectal cancer mortality increased 0.25% annually in the lowest socioeconomic group, while consistently declining in higher socioeconomic groups, reversing the inequality. In the 1950s, mortality was highest in the wealthiest stratum, but by 2009– 2013 the inequality had reversed, with mortality highest in the lowest socioeconomic group^7^.

In Germany, a study found that although incidence declined sharply from 2007 to 2018 for total and most cancers across all social strata, differences in trend magnitudes across deprivation quintiles resulted in widening inequalities over time for total, colorectal, and lung cancers^4^. In developed countries, the highest incidence and mortality rates are recorded among populations with the lowest socioeconomic status^43^.

## Conclusion, Global Context and Broader Implications

Global literature consistently highlights the influence of social inequalities on cancer burden. A scoping review by Costa, Ramos, and Sousa (2024)^44^ on social inequality indicators associated with cancer mortality in Brazilian adults concluded that mortality is influenced by social inequalities. Still, no single association pattern applies to all neoplasms or indicators. The study emphasized the diversity of indicators (such as HDI and Gini coefficient) used to assess income, education, human development, and vulnerability, while pointing out gaps in small-area analyses and the use of multidimensional indicators.

Globally, social vulnerability is a strong predictor of cancer mortality. Chen et al. (2023)^45^ studied U.S. counties and showed that differences in local social vulnerability impact cancer mortality, with the highest rates among the most vulnerable counties. This was especially true among men, non-Hispanic Black individuals, and for lung and colorectal cancers. Using the Social Vulnerability Index, which includes 15 social factors (such as socioeconomic status, family composition, race/language, housing type, and transportation), the study provided a framework for understanding how multiple social determinants translate into health inequities. The pattern documented in Campinas, with higher mortality in vulnerable groups, aligns with this perspective.

A representative population-based cohort study in Canada^46^ found that modifiable risk factors (smoking, excessive alcohol consumption, low fruit/vegetable intake, physical inactivity, and obesity) explained 45.6% of associations between low socioeconomic position and cancer morbidity and mortality. In China, Maomao et al. (2022) estimated 4.8 million new cases and 2.5 million cancer deaths in 2022, with age-standardized incidence rates increasing annually (1.4% between 2000–2018), while age-standardized mortality rates declined (1.3% annually)^47^.

In Poland, the most common cancers were prostate (20.6%), lung (16.1%), and colon (6.8%) in men, and breast (22.9%), lung (9.9%), and uterine corpus (7.0%) in women. Mortality patterns also varied by sex and cancer type, with lung cancer as the leading cause of death in both sexes^48^. Regional and sex-specific variations in Europe^49^ and Asia^50,51,52^ reinforce the need for localized studies such as the one conducted in Campinas to identify specific patterns.

## Policy Implications and Future Directions

The scenario documented in Campinas underscores persistent social inequalities in cancer incidence and mortality, particularly among the most vulnerable populations. This emphasizes the urgency of structured public policies that prioritize equitable access to timely diagnosis, screening, and appropriate treatment. As highlighted by Souza et al. (2023)^53^ and Arık et al. (2021)^21^, the effectiveness of cancer control strategies depends on the health system’s capacity to identify and reach high-risk groups to break cycles of exclusion and mitigate the effects of social inequities on cancer outcomes.

## Data Availability

No ethical and legal restriction for all authors. All data including in this manuscript are public and available in “https://campinas.sp.gov.br/secretaria/saude/pagina/rcbp”

